# Prognostic value of 8q gain in relation to *BAP1* and *SF3B1* mutated uveal melanoma

**DOI:** 10.1101/2022.09.20.22280160

**Authors:** Josephine Q.N. Nguyen, Wojtek Drabarek, Jolanda Vaarwater, Serdar Yavuzyigitoglu, Robert M. Verdijk, Dion Paridaens, Nicole C. Naus, Annelies de Klein, Erwin Brosens, Emine Kiliç, the Rotterdam Ocular Melanoma Study group

**Affiliations:** Department of Ophthalmology, Erasmus MC Cancer Institute, Erasmus MC Medical Center Rotterdam, 3000 CA Rotterdam, The Netherlands; Department of Clinical Genetics, Erasmus MC Cancer Institute, Erasmus MC Medical Center Rotterdam, 3000 CA Rotterdam, The Netherlands; The Rotterdam Eye Hospital, 3011 BH, Rotterdam, The Netherlands; Department of Pathology, Section Ophthalmic Pathology, Erasmus MC Cancer Institute, Erasmus MC Medical Center Rotterdam, 3000 CA Rotterdam, The Netherlands; Department of Pathology, Leiden University Medical Center, 2333 ZA Leiden, The Netherlands

## Abstract

Chromosome 8q gain is associated with poor prognosis. Here, we show that the predictive value of chromosome 8q gain depends on the mutation status and is true for *BAP1* but not for *SF3B1*-mutated tumors.

---

Uveal melanoma (UM) is the most common primary intraocular malignancy in adults [1]. Recurrent mutations in secondary driver genes *BAP1, SF3B1*, and *EIF1AX*, as well as characteristic copy number variations (CNV) and gene expression profiles (GEP) are used in prognostication. Previously, gain of chromosome 8q has been associated with poor prognosis, with increased numbers of 8q correlating with shorter survival [2]. Whereas in *BAP1*-mutated (*BAP1*^MUT^) UM, gain of 8q is the result of whole chromosome 8 gain or the formation of isochromosome 8q, in case of *SF3B1*-mutated (*SF3B1* ^MUT^) UM structural, often partial gain of 8q is predominant [3]. Some of these *SF3B1* ^MUT^ UM patients develop early-onset metastatic disease [4], prompting to investigate the relationship between survival, gain of 8q and *SF3B1* ^MUT^ UM.

Patients with *SF3B1* ^MUT^ tumors (n=59) from the Rotterdam Ocular Melanoma Study group diagnosed between 1994 and 2022 were included in this study. CNVs were assessed using single nucleotide polymorphism (SNP) array (n=55) or in the past with karyotyping (n=17) and or fluorescence in situ hybridization (FISH, n=28) and when material was available used for transcriptome profiling (n=19) [4]. The summary plot showing the chromosomal patterns of the SNP arrays can be found in the supplementary (available at www.aaojournals.org). Disease-free survival (DFS) was determined using a cut-off of 60 months to identify early-onset metastatic disease in UM patients [4].

Gain of chromosome 8q was present in 48 tumors (81%) (Figure 1a). Cox proportional hazard analysis could not confirm reported independent [2] prognostic value of gain of 8q in general (>2 copies) in *SF3B1* ^MUT^ UM (HR: 1.042 (95%;CI:0.3105-4.733)), nor the number of additional 8q copies (3 copies of 8q, HR: 1.213 (95%;CI:0.3363-5.736); ≥4 copies of 8q, HR: 0.6913 (95%;CI:0.08939-4.264)) in *SF3B1* ^MUT^ tumors. Kaplan-Meier survival analysis also indicated no difference in survival in patients with or without 8q gain (*p*=0.9854; Figure 1b) nor is there a difference between 2, 3 or ≥4 copies (*p*=0.6927; Figure 1d). However, since 8q gain is a characteristic of *SF3B1* ^MUT^ as well as *BAP1* ^MUT^ tumors, the prognostic value of 8q gain could also be only attributed to *BAP1* tumors. Therefore, we assessed the survival of 211 UM patients with immunohistochemically BAP1-negative tumors or tumors with *BAP1* mutations. Gain of chromosome 8q was present in 181 tumors (86%) and this was correlated with a worse survival in *BAP1* ^MUT^ UM (*p*=0.0134; Figure 1c). Three copies of 8q was not associated with decreased survival (HR: 1.667 (95%;CI:0.9755-3.042) but more copies (≥ 4 copies) had predictive value (HR: 1.907 (95%;CI:1.103-3.507, Figure 1e). Since (partial) gain of 8q can also be accompanied with changes in 8p copy number, 8p status in *SF3B1* ^MUT^ tumors was also assessed in 8q gain tumors (1 copy 8p (2.1%); 2 copies 8p (77.1%); ≥3 copies 8p (20.8%)) (Figure 1a). Next, the survival plot was stratified on 8p status. Patients with a *SF3B1* tumor and loss or gain of 8p (1 copy 8p, n=1; 3 copies 8q, n=4) in combination with 3 copies of 8q had the shortest survival (*p*=0.0352). Since *BAP1* ^MUT^ tumors have recurrent isochromosome 8q, we explored the combination of 8q and 8p status, where 8q gain was accompanied with 1 copy (40%), 2 copies (35%) and ≥3 copies of 8p (25%). However, no difference was found in 8p status between *SF3B1* ^MUT^ and *BAP1* ^MUT^ tumors (*p*=0.4149). Nonetheless, many aberrations can be overlooked with single probe detection. No difference was found in copies of 8p (*p*=0.2274) and 8q (*p*=0.8237) between DFS<60 months (n=8) and DFS≥60 months (n=51) in *SF3B1* ^MUT^ tumors (Figure 1a). Distal gains (8q23-8q24.3) were similar in patients with a DFS<60 months and DFS≥60 months (71% vs 78%; *p*=0.7122), more proximal (8q1-8q23) gains were more prominent in DFS<60 patients, though this difference was not significant (71% vs 41%; *p*=0.1461). Fifteen patients have metastatic disease (DFS<60 months, n=8 (88% 8q gain); DFS≥60 months, n=7 (71.4% 8q gain)) [4]. When grouping patients based on DFS, the number of 8q copies was not associated with survival (DFS<60, *p*=0.5319; DFS≥60, *p*=0.2328) (Figure 1f-g). Interestingly, ≥4 copies of 8q corresponded to the best survival (100%) in patients with DFS≥60 months. Of the patients with DFS≥60 months and ≥4 copies of 8q, 50% had ≥3 copies of 8p, whereas 25% of the patients with DFS<60 months and ≥4 copies of 8q had ≥3 copies of 8p. Transcriptome profiling and evaluating the genes from the GEP test on *SF3B1* ^MUT^ (n=12) and *BAP1* ^MUT^ (n=7) tumors showed no discriminating factor with 8q status (*p*=0.4149). One of the genes distinguishing DFS<60 and DFS≥60, *TOP1MT* [4] is located at 8q24.3 and did not cluster to any group (Figure 1h).

**Figure 1.**
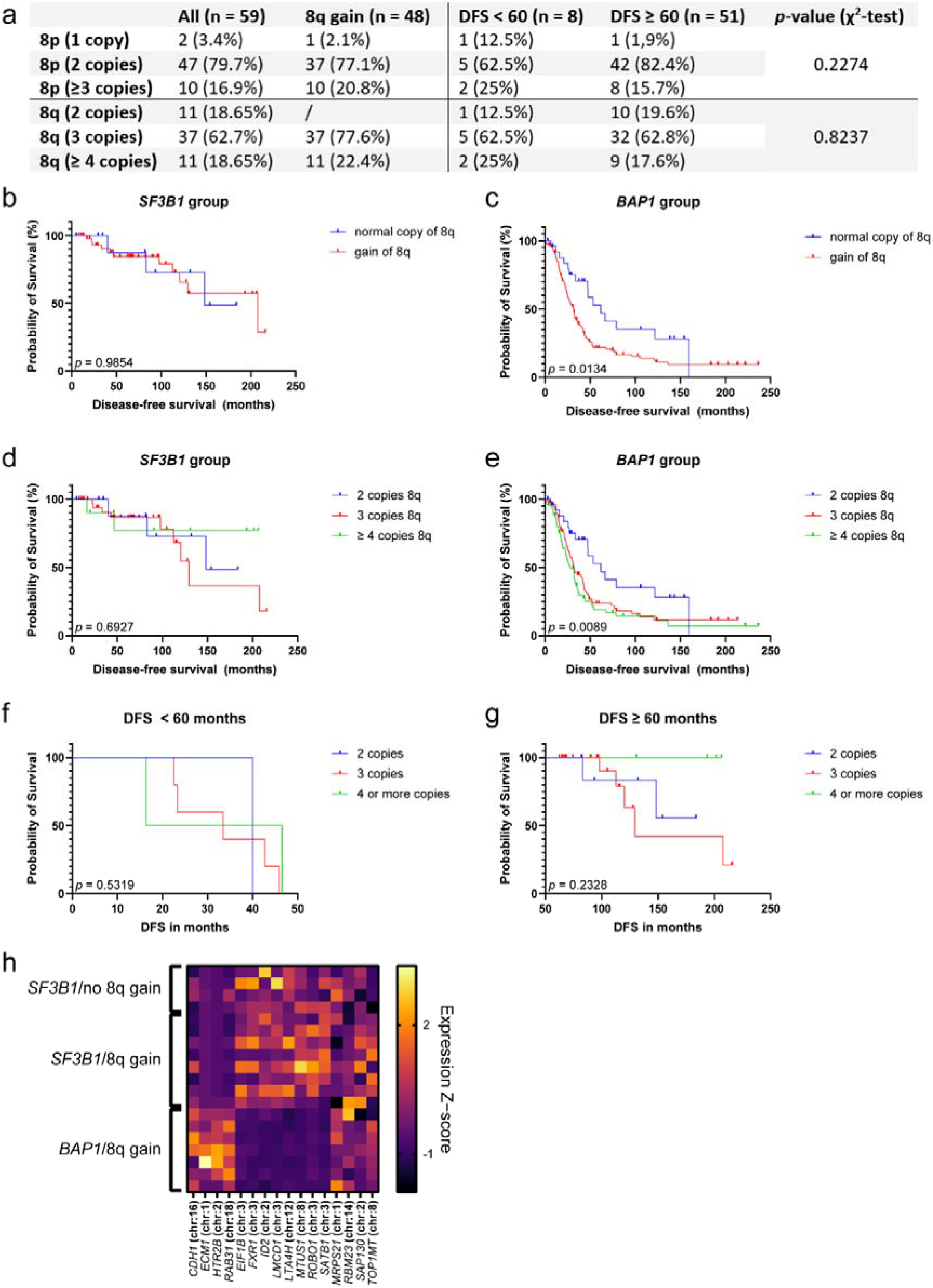
Frequency of 8p and 8q copies in (a) all *SF3B1* ^MUT^ tumors, 8q gain *SF3B1* ^MUT^ tumors, and early (< 60 months) vs late (≥ 60 months) DFS; chromosome 8q status in (b) *SF3B1* ^MUT^ tumors and (c) *BAP1* ^MUT^ tumors; chromosome 8q copy numbers in (d) *SF3B1*^MUT^ tumors and (e) *BAP1*^MUT^ tumors; chromosome 8q copy numbers in (f) early and (g) late DFS *SF3B1*^MUT^ tumors; (h) heatmap using Z-scores of the GEP test of UM including TOP1MT on *SF3B1*^MUT^ and *BAP1*^MUT^ tumors [4]. Samples were clustered based on mutational and 8q status. No *BAP1*^MUT^ tumors had normal copies of 8q.

*SF3B1* ^MUT^ and *BAP1* ^MUT^ tumors often have (partial) 8q gain [3, 5]. Gain of 8q is correlated to prognosis in UM, with increased metastatic risk correlates with increased 8q copy numbers, often a result of isochromosome formation [2, 5]. Here, we show that 8q gain has no discriminating association with survival in early versus late metastasizing *SF3B1* ^MUT^ tumors. Nevertheless, gain of 8q could still play a role in metastases in *SF3B1* ^MUT^ tumors, independent of DFS. *BAP1* ^MUT^ tumors contribute to >50% of all UM and are characterized by gains and losses of entire chromosomes or chromosome arms. Gain of 8q is often accompanied with monosomy 3, and its combination correlates with a worse prognosis [6, 7]. In contrast, *SF3B1* ^MUT^ UM karyotypes are more complex and CNV often have recurrent distal breakpoints on chromosome 6 and 8. This difference in CNV patterns could indicate a separate tumorigenesis mechanism in both groups. Since 8q gain was correlated with survival in only *BAP1* ^MUT^ tumors and not *SF3B1* ^MUT^ tumors, the reported decreased survival in 8q gain is predominantly due to *BAP1* ^MUT^ tumors.

To conclude, we have evaluated gain of chromosome 8q and its role on DFS in *SF3B1* ^MUT^ and *BAP1* ^MUT^ UM. There is no correlation between 8q gain and early-onset metastasis in *SF3B1* ^MUT^ tumors. Gain of 8q has no additional predictive value in *SF3B1* ^MUT^ tumors. In contrast, 8q gain is predictive for a worse prognosis in *BAP1* ^MUT^ tumors. Thus, 8q gain has additional predictive value for *BAP1* ^MUT^ tumors, but not for *SF3B1* ^MUT^ tumors.

## Data Availability

All data produced in the present study are available upon reasonable request to the authors

