## Supplementary figures and images for "Prognostic value of 8q gain in relation to *BAP1* and *SF3B1* mutated uveal melanoma"

### Supplementary figure 1

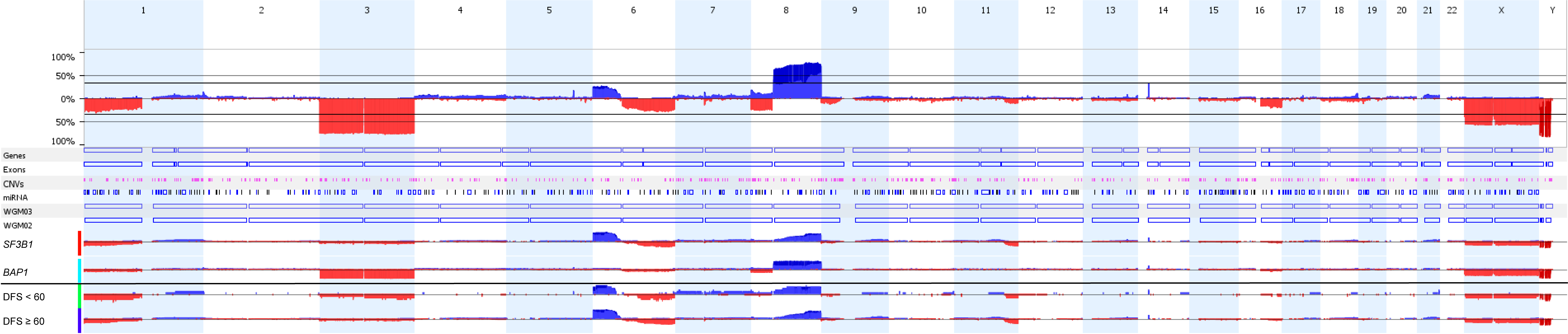
